# Modelling the impact of the El Niño–Southern Oscillation (ENSO) on the dynamics of Dengue outbreaks in Argentina between 2018 and 2024

**DOI:** 10.64898/2026.09.03.26362163

**Authors:** Chiara Elizabeth Lodigiani, Alejandro Enet, Maia Arrúa Miano, Gabriela Lakkis, Lucas Mariano Moreno Lell, Carla María Panzarella, Sol Denise Otero Legovich, Ornella Abril Bellizzi, Manuela Bulló, Santiago Perez-Lloret

## Abstract

**Background:** Dengue fever has become an epidemiological concern worldwide, with 531,617 cases reported in Argentina during the 2023-2024 season. The El Niño– Southern Oscillation (ENSO) phenomenon has been linked to changes in dengue dynamics. This study assessed the relationships among ENSO, local climatic variables, and dengue outbreaks in Argentina from 2018 to 2024.

**Methods:** A nationwide spatiotemporal analysis was conducted using dengue cases and monthly mean temperature, relative humidity, and rainfall. The Southern Oscillation Index (SOI) was used as the ENSO indicator (El Niño phase=< −0.5, neutral phase= −0.5 to 0.5, and La Niña phase= > 0.5). We fitted Hierarchical Bayesian models with Integrated Nested Laplace Approximation (INLA). A Negative Binomial distribution of dengue cases was used to account for overdispersion. Spatial dependence was modeled by using the Besag–York–Mollié 2 (BYM2) structure, adjusted for population.

**Results:** The final model explained 53.7% of the variation in Argentina’s departmental monthly dengue cases during the study period. Exposure-response curves of posterior probability and 95% credible intervals suggested that dengue risk increased when monthly mean temperatures were between 20° and 30° Celsius during the El Niño or neutral phase, with less consistent effects during the La Niña phase. Humidity >50% was associated with increased dengue risk during the La Niña phase, with less steep or absent associations during El Niño or neutral phases. Negative SOI scores were associated with an increased risk of dengue after controlling for the covariates.

**Conclusion:** Dengue outbreaks in Argentina were independently associated with temperatures between 20 and 30 °C during the El Niño and neutral phases, humidity > 50% during the La Niña phase, and the El Niño phase of the ENSO. Incorporating ENSO indices into predictive models could enhance early warning systems and timely public health interventions in Argentina.

**HIGHLIGHTS:**

- This study linked the El Niño phenomenon to Dengue fever outbreaks in Argentina.
- Other predictors of increased risk were monthly mean temperatures between 20° and 30° Celsius during the El Niño and neutral phases, and humidity > 50% during the La Niña phase.
- There was a high degree of spatial residual variance, not accounted for by the factors considered in the model, in dengue risk, which was increased in some northern and southern provinces of Argentina.

## 1. INTRODUCTION

Dengue is a viral disease transmitted by mosquitoes of the genus *Aedes*, mainly *Aedes aegypti* (Jansen, C.C., 2010; Roy S.K., 2021). The causative agent is the dengue virus, which belongs to the Flaviviridae family and has four different serotypes (DENV-1, DENV-2, DENV-3, and DENV-4) (Roy, S. K., 2021). Dengue is endemic in many tropical and subtropical regions worldwide, with higher incidence during rainy seasons, when mosquito populations increase (Roy, S. K., 2021). Approximately 380 million cases and 22,000 deaths occur each year. In Argentina, 531,617 cases were registered during the 2023-2024 season with 419 deaths (Kantor I. N., 2024). The most affected regions were the northern provinces.

Several studies in tropical and subtropical regions of Latin America have demonstrated a significant correlation between dengue incidence and climatic variables, with temperature, humidity, and, to a lesser extent, rainfall as the primary factors (López et al., 2023; Ferreira H. D. S., 2022). There has been a significant increase in Earth’s temperature of about 1.16 Celsius degrees since 1951 (Wang L., 2023), which may have contributed to the acceleration of the dengue burden (López et al., 2023).

Higher temperatures accelerate dengue virus transmission by reducing the vector incubation period, increasing the contagion rate, and causing an exponential increase in cases (De Almeida M. T., 2025). Rainfall and ambient humidity improve vector survival rates by promoting the accumulation of stagnant water, which supports mosquito development. (Ferreira H. D. S., 2022; Li C., 2021).

The El Niño–Southern Oscillation (ENSO) is a recurring climate pattern that results from variations in ocean temperatures in the central and eastern equatorial Pacific Ocean, coupled with changes in atmospheric pressure patterns across the tropical Pacific (Gagnon et al., 2001; McPhaden et al., 2006; Ropelewski, C. F., 1987). ENSO has two main phases: El Niño (the warm phase) and La Niña (the cool phase), as well as a neutral phase (Trenberth, K.E., 1997). During El Niño events, abnormally warm sea surface temperatures develop in the eastern Pacific, disrupting typical weather patterns and leading to increased rainfall and humidity in some regions, while causing droughts in others. (Mokhtar et al., 2024) Conversely, La Niña events are characterized by cooler-than-average sea surface temperatures in the same region, which strengthens the trade winds and often results in opposite weather anomalies-drier conditions in some areas and increased rainfall in others. In Argentina, the El Niños phase is associated with increased ambient humidity, and the La Niña phase with dry conditions.

Several studies suggest a relationship between ENSO and dengue outbreaks. A recent study found that El Niño was positively correlated with the global risk of dengue outbreaks, with a correlation coefficient of 0.52 (Mokhtar, S., 2024). These results have been replicated in some regions of Brazil. A study conducted across 645 municipalities in the state of São Paulo (Brazil) linked El Niño events to increases in the mosquito larval index, a measure of mosquito proliferation (Pirani, M., 2024).

In this study, we aimed at modeling the influence of ENSO on dengue outbreaks in Argentina, accounting for the effects of local climate parameters.

## 2. METHODS

### 2.1 Data collection

We conducted a nationwide analysis of dengue case distribution in Argentina between 2018 and 2024 at a monthly resolution. Argentina is in the southern part of South America, covers an area of 2,780,400 km² of continental and insular territory, and has a population of just over 46 million inhabitants. Argentina stretches over 3,700 km from north to south, which explains why it spans a remarkable variety of climates, from subtropical in the north to subpolar in the far south (Prohaska, F., 1976). This great length also gives rise to diverse biomes, including rainforests, grasslands, deserts, and Patagonian tundra. The country is divided into 525 departments and an autonomous city, Buenos Aires (hereinafter referred to as departments). Data on dengue incidence were obtained from the National Ministry of Health Surveillance System 2.0, accessed through https://datos.salud.gob.ar/dataset/vigilancia-de-dengue-y-zika (Date of Accession: 2025-02-01). A dengue case was defined as any confirmed or probable dengue infection reported through the national health surveillance system. For each department, monthly dengue incidence was calculated as the number of cases per 100,000 inhabitants. Population and population density data for each department were sourced from the 2022 National Census conducted by the National Institute of Statistics and Censuses (INDEC, https://www.indec.gob.ar/, Date of Accession: 2025-02-01).

Climatic data were obtained from NASA’s POWER Project (Prediction of Worldwide Energy Resources) (Zhang, T., 2007). For each department and month, we used the monthly mean of daily values for temperature (measured in degrees Celsius, °C) and relative humidity (measured in %), both at 2 meters above ground, as well as the monthly accumulated rainfall (measured in millimeters, mm). The spatial resolution of the satellite data is 0.5° latitude × 0.625° longitude (approximately 50 km). Daily values were extracted for each centroid within each administrative department and subsequently averaged to obtain monthly means; for rainfall, daily values were summed to derive monthly totals.

The Southern Oscillation Index (SOI) was also considered as a measure of ENSO (Tipayamongkholgul et al., 2009; Trenberth, K.E., 1984). This index reflects differences in atmospheric pressure between Tahiti and Darwin, Australia, and is used to characterize the El Niño and La Niña phases. SOI scores were used in the analyses as a measure of the magnitude and direction of ENSO variability, which facilitates the climatological interpretation of exposure–response relationships. SOI scores above 0.5 indicated the La Niña phase; scores below −0.5 indicated the El Niño phase, with values between −0.5 and 0.5 characterizing a neutral phase, following the threshold criteria used in previous ENSO dengue studies (Pirani M., 2024).

### 2.2 Statistical analysis

We fitted hierarchical Bayesian models using the Integrated Nested Laplace Approximation (INLA) framework in R 4.5 (R Foundation for Statistical Computing, Vienna, Austria; R-INLA package).

The response variable was the monthly number of dengue cases, and we assumed a Negative Binomial likelihood to account for overdispersion.

The general model fitted was:

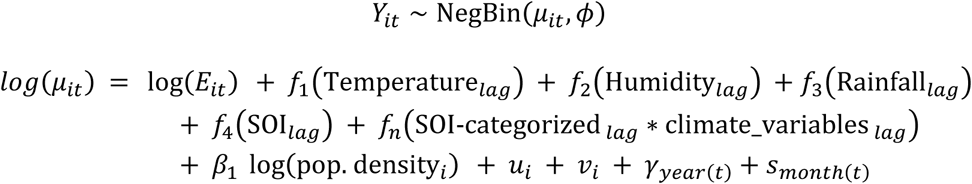

Where Yit denotes the number of dengue cases observed in department i during month t, assumed to follow a negative binomial distribution. The term log(Eit) denotes the logarithm of the population size and is used as an offset. Nonlinear effects of temperature, relative humidity, rainfall, and SOI, as well as their interactions, were modeled using second-order random walks (RW2), which impose smoothness by penalizing second differences between adjacent latent effects. The RW2 terms were fitted using the default INLA penalized complexity priors for the precision parameter. For the RW2 components, climate parameters and SOI were centered and transformed into ordered indices representing adjacent exposure levels. Temperature and relative humidity were rounded to the nearest integer and converted to sequential indices, rainfall was rounded to the nearest 5-mm interval and converted to sequential indices, and SOI anomalies were rounded to the nearest integer and indexed similarly. These indices were then used as the covariates in the RW2 smooth terms, allowing estimation of nonlinear exposure–response relationships across the observed range of values. Population density was log-transformed and standardized before being included as a linear fixed effect.

Interactions between the ENSO phase and climatic variables were implemented using the replicate argument in INLA. Specifically, separate RW2 smooth functions were estimated for temperature and relative humidity within each SOI category (El Niño, Neutral, and La Niña), while sharing a common smoothing prior structure. The terms *ui* and *vi* represent the structured and unstructured spatial random effects, respectively, modeled using the Besag–York–Mollié 2 specification. Departmental neighborhood relationships were defined using a first-order contiguity adjacency matrix derived from the departmental polygon shapefile, in which two departments were considered neighbors if they shared a common boundary. This adjacency matrix was supplied to INLA through the BYM2 specification. The BYM2 spatial effect was fitted using the default penalized complexity (PC) priors implemented in R-INLA.

Temporal dependence was modeled through a 12-month cyclic seasonal effect to account for recurrent intra-annual patterns, together with an independent random effect for year to capture interannual variability in dengue transmission.

We explored the best-fitting lags for SOI and climate parameters by computing and comparing the Deviance Information Criterion (DIC) and the Widely Applicable Information Criterion (WAIC) between the models. Candidate lags from 0 to 11 months were evaluated independently for each climatic variable and SOI. The lag yielding the lowest DIC and WAIC values was retained in the final model.

The interactions between the SOI and the climatic variables at their best-fitting lags were explored by categorizing SOI values, as established in the literat into El Niño (<-0.5), Neutral (−0.5 to 0.5), and La Niña (> 0.5) phases (Pirani, M., 2024). Models were fitted, and DIC and WAIC were used to select the best model.

The final model adequacy was assessed through posterior predictive checks, including the randomized Probability Integral Transform (PIT) histogram, Q–Q plots of the randomized PIT values, and simulation-based residual diagnostics using the DHARMa package. DHARMa diagnostics included quantile residual Q–Q plots and residual-versus-predicted value assessments to evaluate potential deviations from distributional assumptions, overdispersion, and residual structure. To assess model performance, we calculated a pseudo-R² using Nagelkerke’s method, based on the deviance values from the fitted and null models. Final inferences are presented as posterior means and 95% credible intervals from the INLA posterior summaries. Exposure-response curves were then obtained. When interactions with SOI were present, exposure–response curves had three curves, one for each SOI category.

## 3. RESULTS

The total number of dengue cases reported from 2018 to 2024 was 1,559, 2,754, 52,027, 3,852, 793, 139,525, and 492,287, respectively (Figure 1A). No major differences in mean temperature or relative humidity were observed between 2018 and 2024 (Figure 1B–C). In contrast, the Southern Oscillation Index (SOI) exhibited marked temporal variability, with predominantly negative values between 2018 and 2020, followed by positive values from 2020 to 2023, and a decline thereafter.

**Figure 1.**
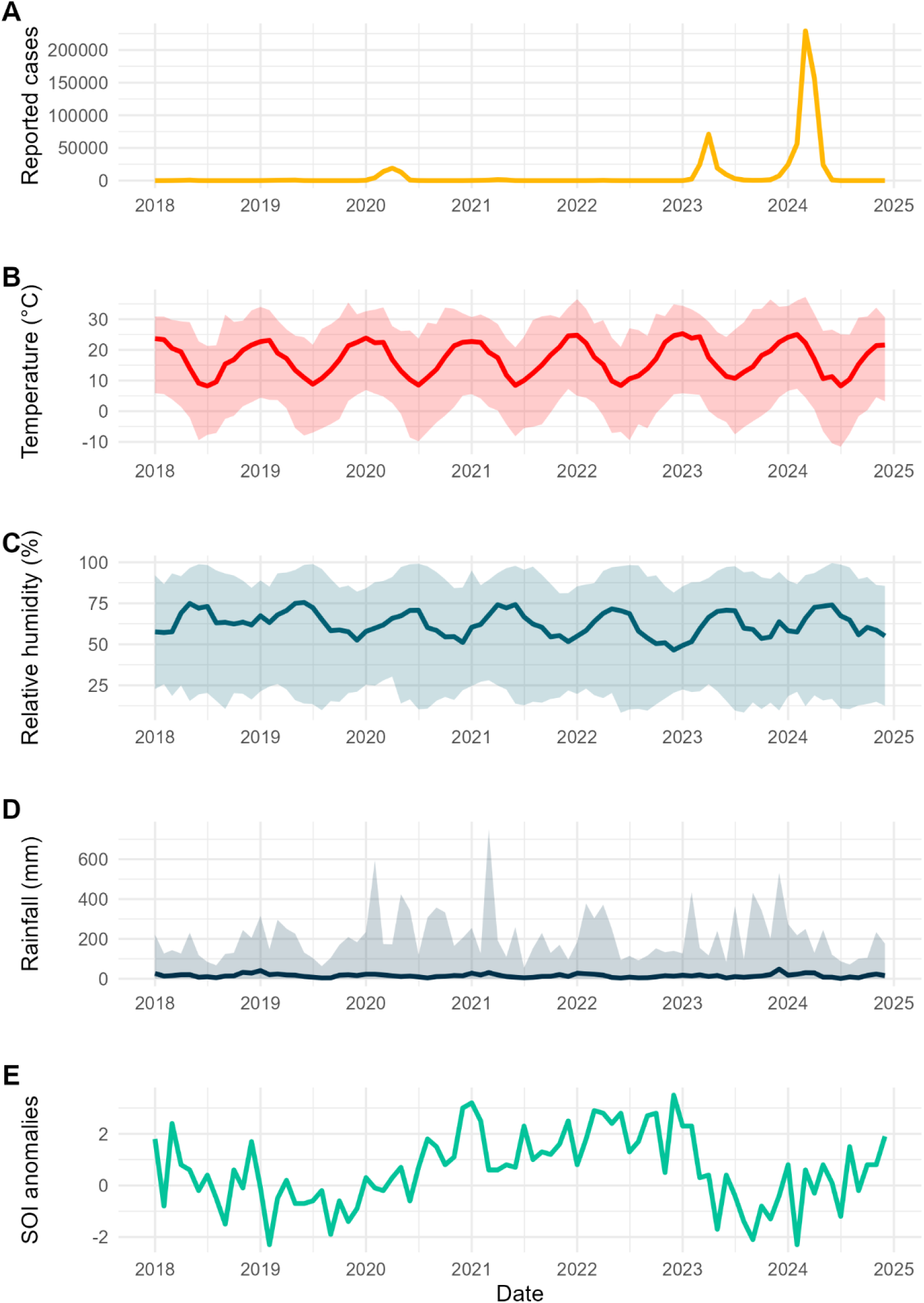
Temporal Patterns of Dengue Incidence and Environmental Variables in Argentina (2018-2024). Shaded areas represent the maximum and minimum values for climate parameters across the country.

Table 1 shows the fit of models involving different lags for the climatic and SOI variables. The best-fitting lag for temperature and humidity was 2 months, for rainfall was 3 months, and for SOI, 5 months. The final model included the variables at the best-fitting lags. Table 2 compares the fit of the different models tested. The introduction of SOI-climate parameter interaction terms reduced DIC and WAIC values and increased Nagelkerke R^2^. When the SOI*temperature and SOI*humidity terms were considered together, the fitting measures improved. Conversely, when the SOI*precipitation term was added, WAIC increased considerably. Therefore, the model retained contained SOI*temperature and SOI*humidity interaction terms plus the main effect terms for all other variables.

**Table 1.** Identification of the best lags in climatic and SOI variables included in the model.

| Lag (months) | Temperature | Humidity | Rainfall | SOI |
| --- | --- | --- | --- | --- |
| 0 | 56566 / 56733 | 56568 / 56734 | 56568 / 56735 | 56565 / 56731 |
| 1 | 56321 / 56502 | 56480 / 56653 | 56562 / 56734 | 56332 / 56509 |
| 2 | **56138 / 56332** | **56265 / 56483** | 56368 / 56551 | 56624 / 56794 |
| 3 | 56427 / 56639 | 56496 / 56682 | **56366 / 56548** | 56281 / 56540 |
| 4 | 56326 / 56545 | 56546 / 56763 | 56425 / 56610 | 56215 / 56393 |
| 5 | 56369 / 56540 | 56666 / 56873 | 56540 / 56705 | **55929 / 56182** |
| 6 | 56279 / 56453 | 56636 / 56798 | 56520 / 56700 | 56621 / 56796 |
| 7 | 56349 / 56532 | 56576 / 56754 | 56431 / 56614 | 56251 / 56457 |
| 8 | 56263 / 56500 | 56444 / 56612 | 56455 / 56633 | 56593 / 56774 |
| 9 | 56197 / 56422 | 56206 / 56406 | 56493 / 56664 | 56653 / 56818 |
| 10 | 56202 / 56412 | 56278 / 56598 | 56435 / 56603 | 56766 / 56948 |
| 11 | 56208 / 56429 | 56290 / 56494 | 56388 / 56567 | 56761 / 56930 |

DIC / WAIC values are shown for each variable at each lag. The minimum values, highlighting the best-fitting lags, are marked in bold.

**Table 2.** Exploration of models with principal effect terms and interactions.

| Model | DIC | WAIC | Nagelkerke R<sup>2</sup> |
| --- | --- | --- | --- |
| Base model: SOI + Temperature + Relative Humidity + Rainfall + Pop. Density | 55874 | 56073 | 0.512 |
| Base model + Temperature * SOI | 53514 | 53696 | 0.533 |
| Base model + Humidity* SOI | 53671 | 53464 | 0.533 |
| Base model + Rainfall* SOI | 53734 | 53568 | 0.532 |
| Base model + Temperature * SOI + Humidity* SOI | 53223 | 53463 | 0.537 |
| Base model + Temperature * SOI + Humidity* SOI + Rainfall * SOI | 53226 | 62455 | 0.537 |

The final selected model was a Bayesian hierarchical negative binomial model in which temperature and relative humidity were modeled as nonlinear effects with a 2-month lag, rainfall as an effect with a 3-month lag, and SOI anomalies score as an effect with a 5-month lag, with interactions between SOI anomalies and both temperature and relative humidity. A complete summary of the final model specification is provided in Table 3. The final model equation is:

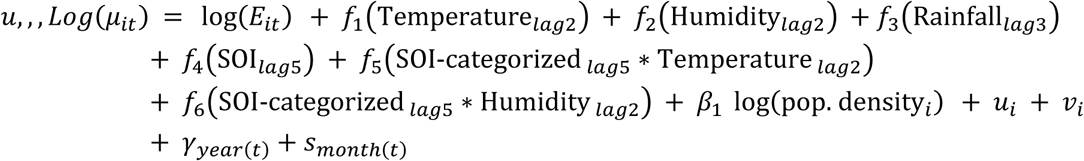

**Table 3.** Summary of the final Bayesian spatiotemporal model.

| Model component | Specification | Significance |
| --- | --- | --- |
| Likelihood | Negative binomial | Compensation for overdispersed counts |
| Response | Monthly dengue counts | Study outcome |
| Spatial effect | BYM2 | Spatial structured and unstructured heterogeneity |
| Temporal effect | Seasonal (month) + IID year | Controls for seasonality and year-to-year variation. |
| Temperature | RW2, lag 2 | Nonlinear main effect |
| Humidity | RW2, lag 2 | Nonlinear main effect |
| Rainfall | RW2, lag 3 | Nonlinear main effect |
| SOI score | RW2, lag 5 | Nonlinear main effect |
| Temperature × SOI-categorical | RW2 replicated by ENSO phase | Nonlinear interaction term |
| Humidity × SOI-categorical | RW2 replicated by ENSO phase | Nonlinear interaction term |
| Population density | Linear standardized effect | Linear main effect |
| Offset | log(population) | Incidence adjustment |

BYM2: Besag–York–Mollié 2 structure; ENSO: El Niño–Southern Oscillation; SOI: Southern Oscillation Index; RW2: second-order random walk; AR1: first-order autoregressive process.

Exposure-response curves for posterior estimates of dengue relative risk across the predictors, along with the corresponding 95% credible intervals, are shown in Figure 2. Temperature (Figure 2A) was a risk factor when the monthly means were between 20 and 30 °C for the El Niño and neutral phases. During the El Niño phase, a monthly mean temperature of 25°C was associated with an estimated RR of 2.92 (95% CrI: 1.69–4.23). Above 30°C, the estimated risk declined markedly. Credible intervals were wider for the La Niña phase, signaling less consistent effects. Humidity (Figure 2B) showed a strong relationship with dengue risk during the La Niña phase, with values exceeding 50% indicating a higher risk. At a monthly mean relative humidity of 70%, the estimated log RR was 2.31 (95% CrI: 1.78–2.92). This relationship was less steep or absent in the El Niño and neutral phases. Precipitation (Figure 2C) showed no clear association with dengue risk. The SOI score showed an inverse relationship with dengue risk after adjusting for the covariates (Figure 2D). Finally, population density (Figure 2E) was a significant predictor of dengue risk.

**Figure 2.**
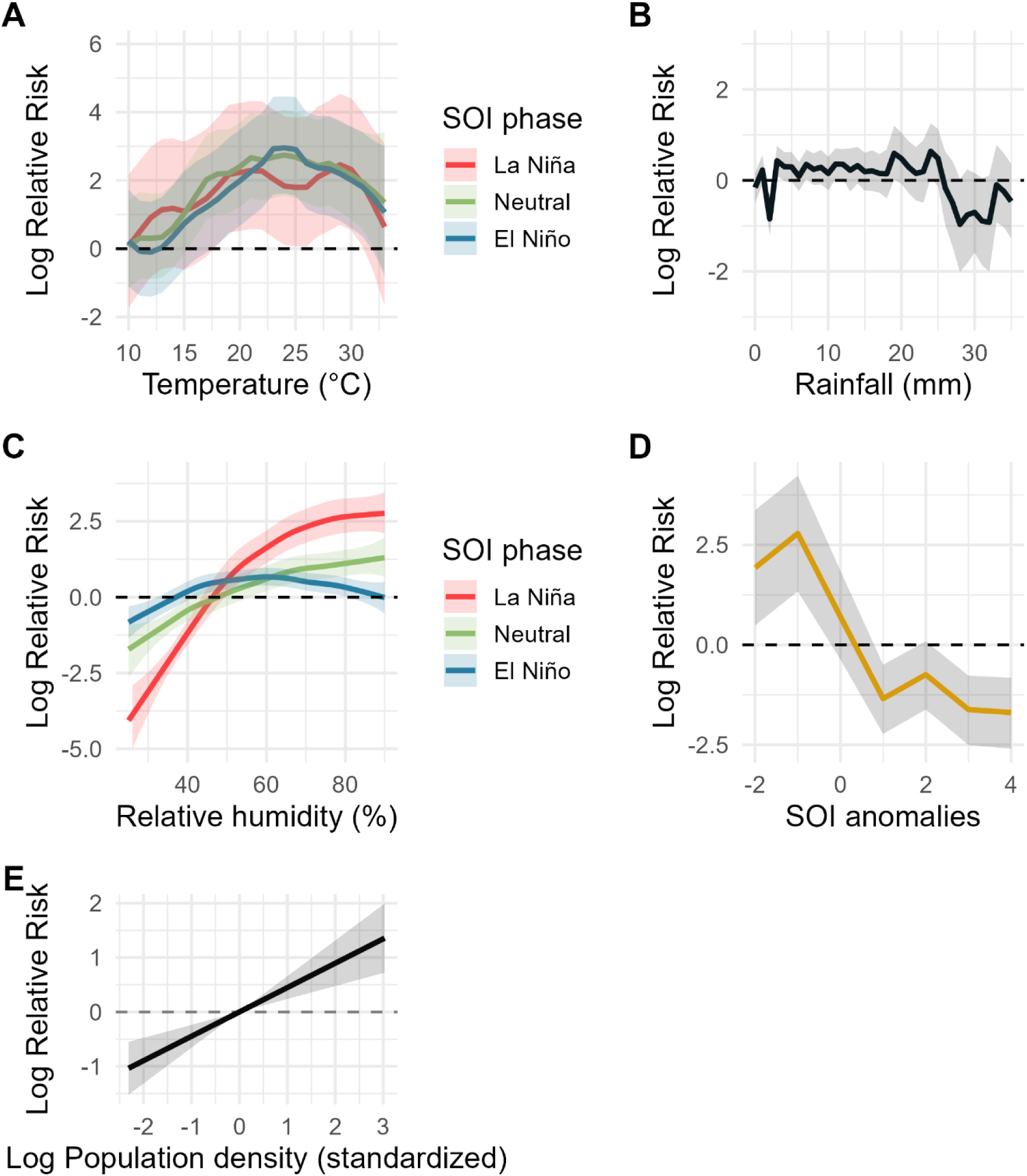
Exposure-response curves for posterior estimates of dengue risk across the predictors, along with the corresponding 95% credible intervals. Panels A–D show the nonlinear associations between dengue risk and (A) temperature (2-month lag), (B) humidity (2-month lag), (C) rainfall (3-month lag), and (D) SOI (5-month lag), modeled using second-order random walks (RW2). Panel E shows the linear effect of standardized log population density. Solid lines represent posterior mean log-relative risks, and shaded areas indicate 95% credible intervals. Values above zero indicate an increased dengue risk relative to the reference level, whereas values below zero indicate a decreased risk.

Figure 3 shows the residual spatial variation captured by the BYM2 component after accounting for climatic, temporal, and demographic covariates. Positive residual spatial effects were concentrated in north-eastern departments, indicating higher dengue risk than expected based on the variables included in the model, whereas many central regions showed lower-than-expected risk. Elevated residual effects were also observed in some southern departments, suggesting that additional unmeasured spatial factors may contribute to the observed geographic distribution of dengue incidence.

**Figure 3.**
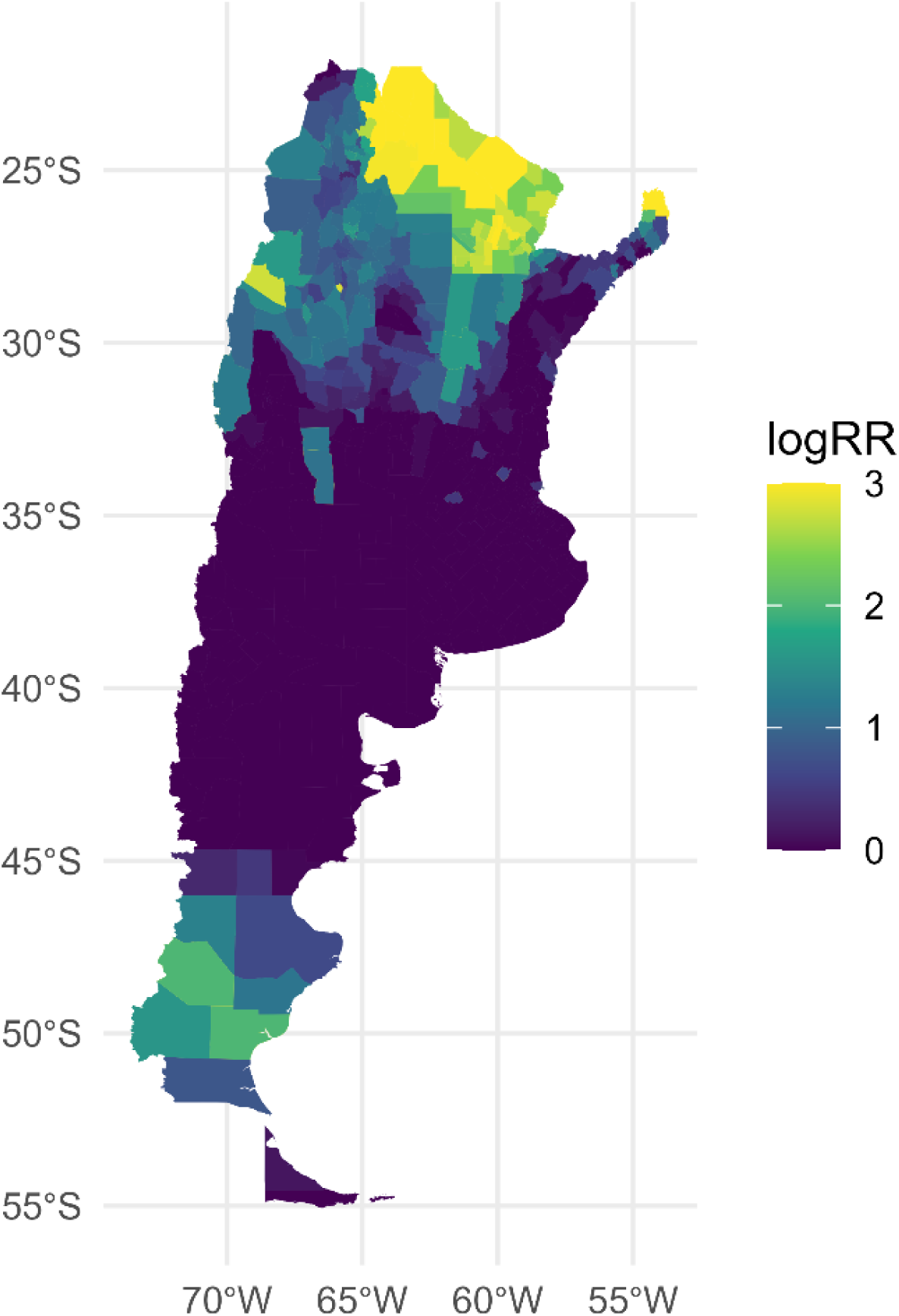
Spatial distribution of the estimated department-level random effects from the Bayesian spatiotemporal model. Colors represent the posterior mean of the spatial log-relative risk derived from the Besag–York–Mollié 2 (BYM2) spatial component. These values reflect residual spatial variation after accounting for climatic, temporal, and demographic covariates included in the model. Positive values indicate departments with higher-than-expected dengue risk, whereas negative values indicate lower-than-expected risk. This figure represents the residual spatial component of the model and should not be interpreted as a map of predicted dengue incidence.

Model diagnostics indicated an adequate fit of the final model. The randomized PIT histogram showed an approximately uniform distribution, and randomized PIT quantiles closely followed the expected diagonal line (Figure 4). Likewise, DHARMa residual diagnostics did not reveal major deviations or systematic residual patterns, supporting the adequacy of the model specification (Figure 5).

**Figure 4.**
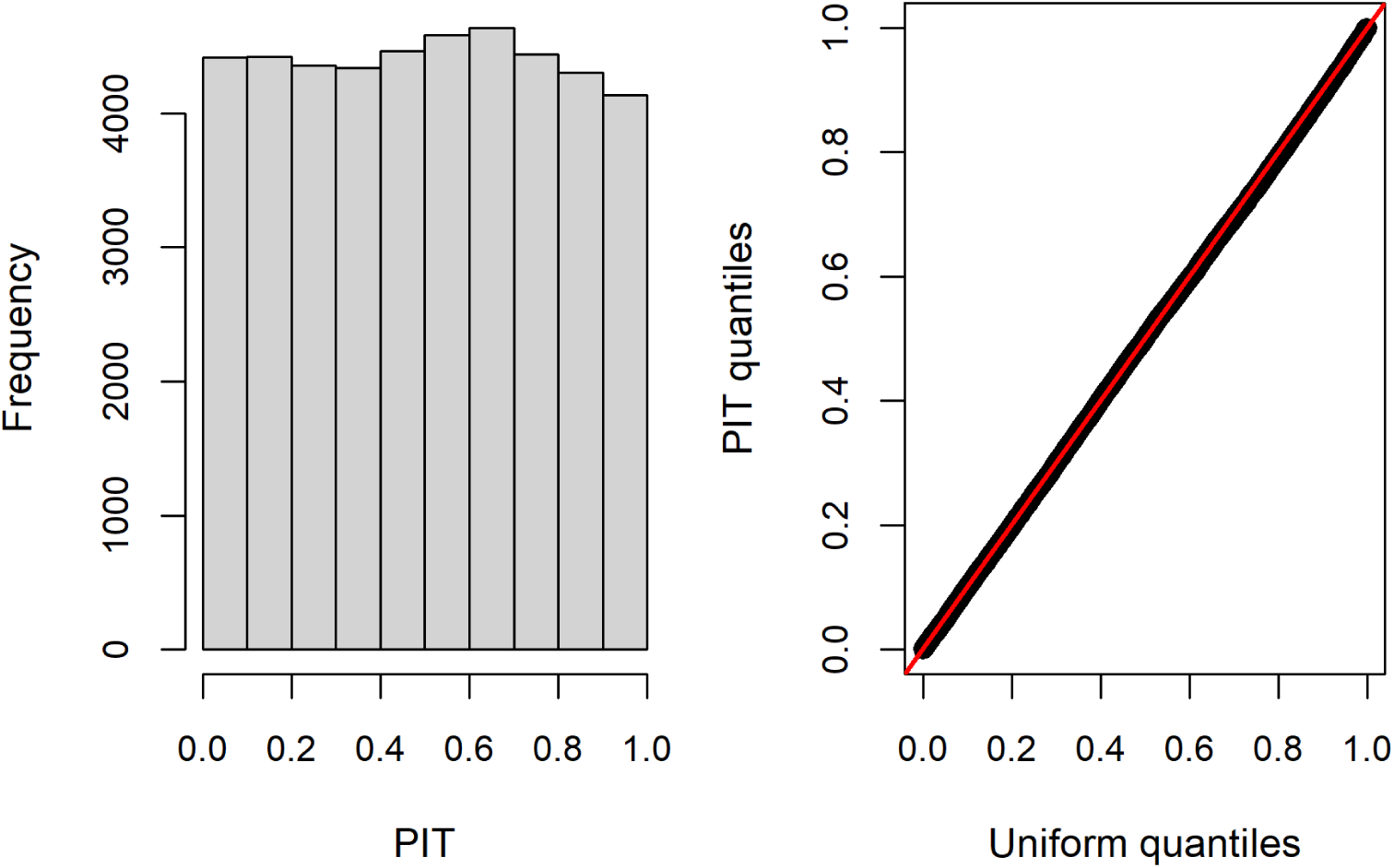
Posterior predictive diagnostics for the final Bayesian spatiotemporal negative binomial model. The left panel shows the histogram of randomized Probability Integral Transform (PIT) values, while the right panel presents the Q–Q plot comparing PIT quantiles against the theoretical uniform distribution. The approximately uniform PIT distribution and close agreement with the reference line indicate adequate model calibration and overall goodness-of-fit.

**Figure 5.**
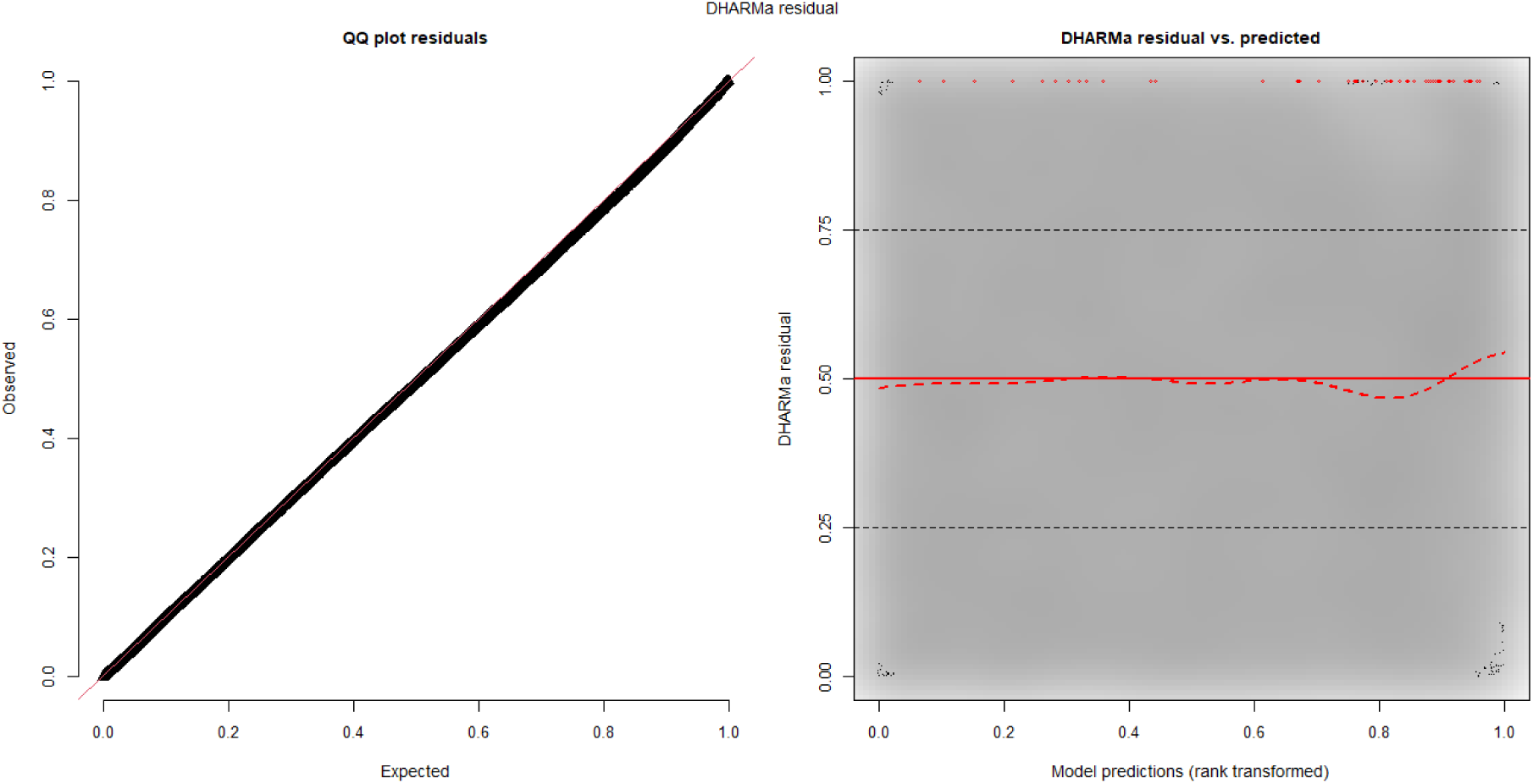
Simulation-based residual diagnostics from the DHARMa package for the final Bayesian spatiotemporal negative binomial model. The left panel shows the Q–Q plot of simulated residuals against the expected uniform distribution, while the right panel presents residuals versus fitted values. The absence of strong systematic deviations and the approximately uniform residual distribution suggest an adequate overall model fit.

Sensitivity analyses further supported the robustness of the selected model. There were no improvements in DIC and WAIC after removing interaction terms from the model (Table 4, rows 1-3), after changing the scale or lag for the SOI (rows 4-8), or when using alternative temporal structures (rows 9-10). The direction and interpretation of the SOI effects were stable across the alternative models evaluated, except for row 7 (SOI as a continuous variable with a 6-month lag).

**Table 4.**
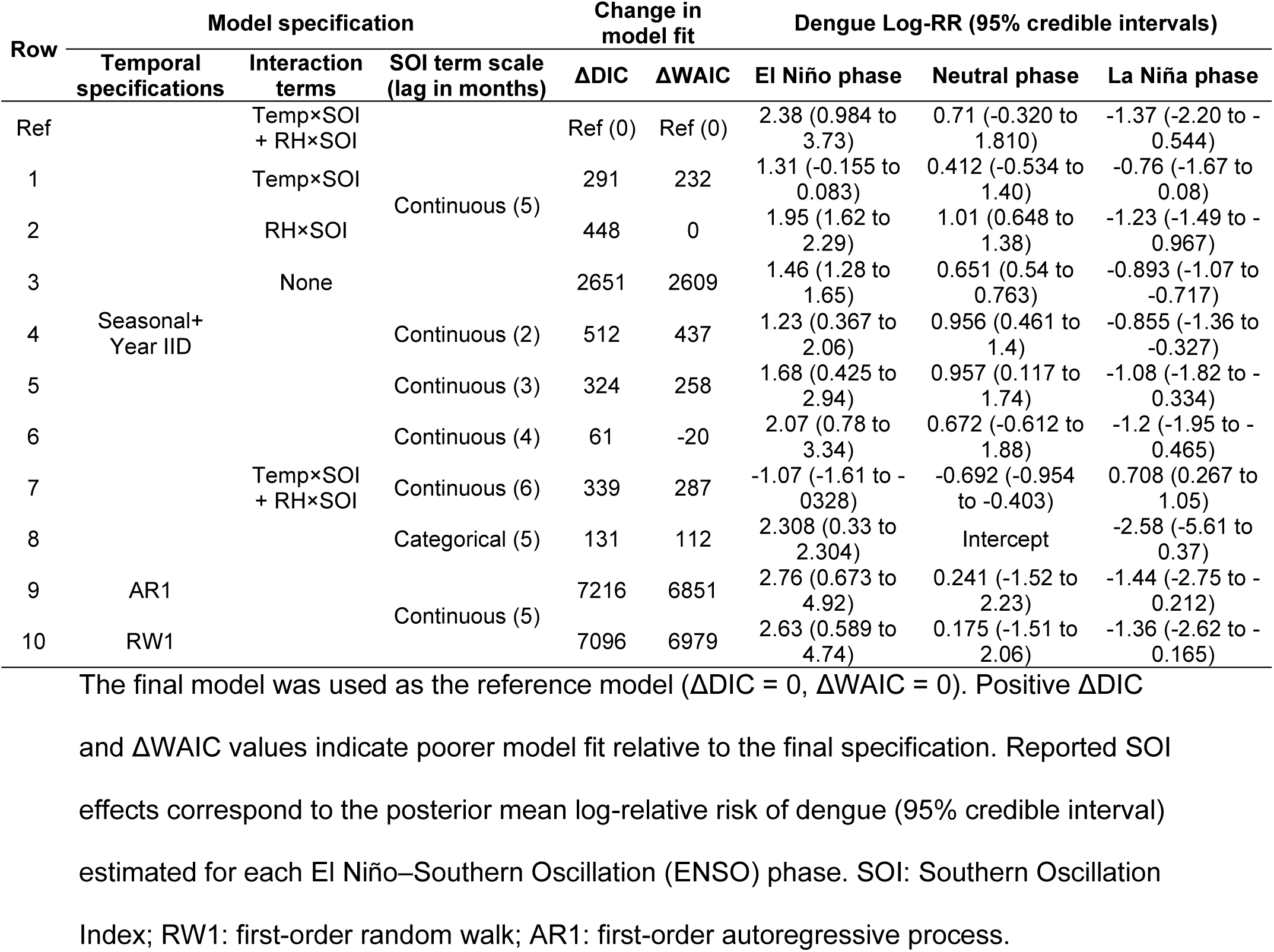
Sensitivity analyses evaluating the model fit and the robustness of SOI effect estimates under alternative model specifications.

Assessment of dependence among the predictors revealed weak correlations among the climatic covariates (Figure 6). The highest correlation was observed between humidity and rainfall (r = 0.40), whereas SOI showed minimal correlation with local meteorological variables (|r| ≤ 0.07). Variance inflation factors were 1.34 for temperature, 1.52 for humidity, 1.44 for rainfall, 1.00 for SOI, and 1.11 for population density, indicating no evidence of problematic multicollinearity among predictors.

**Figure 6.**
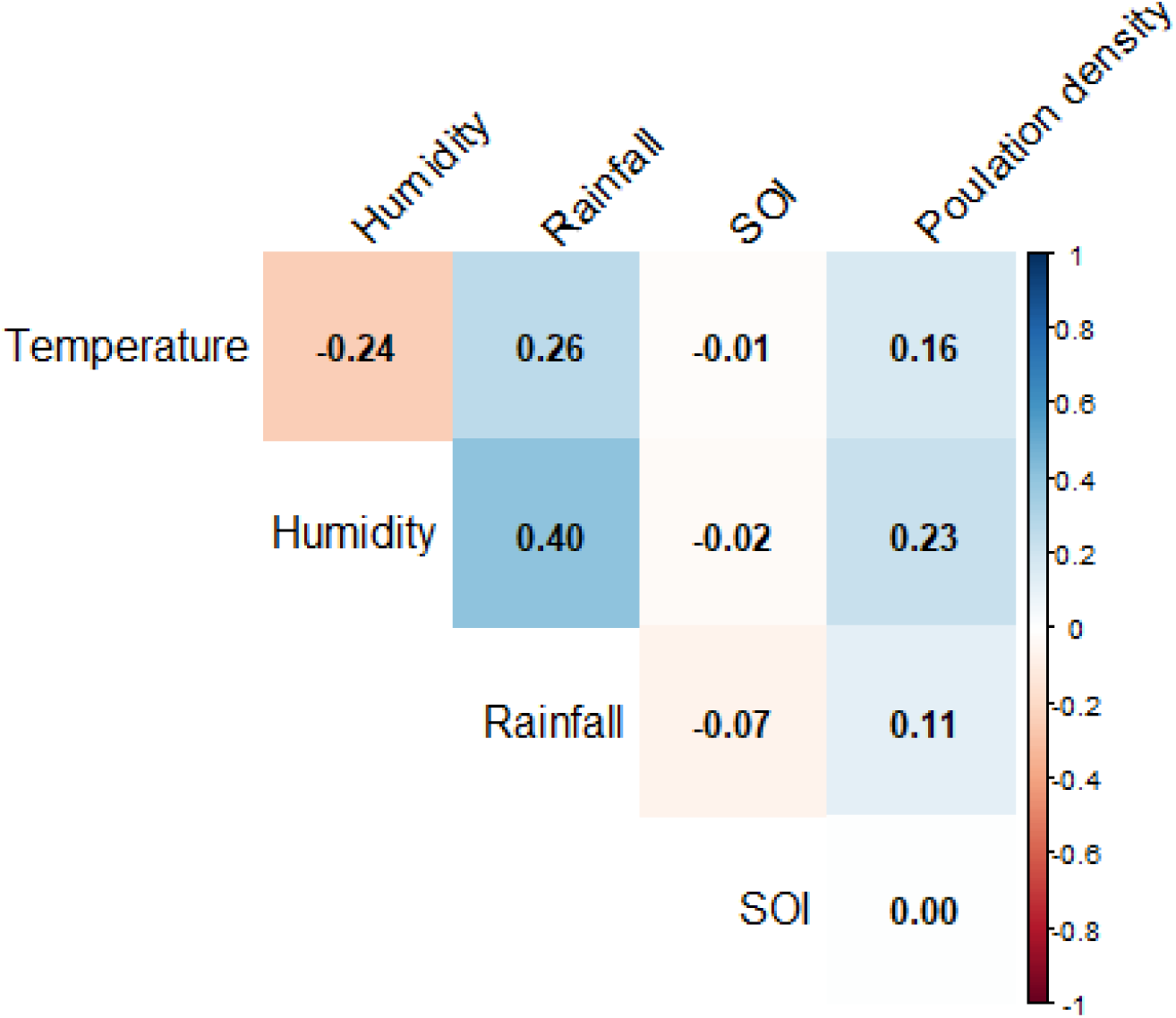
Correlation matrix of the covariates included in the final Bayesian spatiotemporal model. Colors indicate the strength and direction of pairwise Pearson correlation coefficients among temperature, relative humidity, rainfall, SOI anomalies, and standardized log population density. Numerical values represent correlation coefficients. Overall, correlations were weak, with the highest correlation observed between relative humidity and rainfall (r = 0.40, corresponding to R^2^= 0.16), indicating no evidence of substantial collinearity among predictors.

## 4. DISCUSSION

Our results suggest a close relationship between dengue outbreaks in Argentina between 2018 and 2024 and the El Niño phenomenon. Such a relationship has been observed in other parts of the world. In Asia, a study conducted in Thailand between 1996 and 2005 examined the impact of ENSO on dengue outbreaks in two geographically contrasting regions: a tropical coastal region in the south and a mountainous region in the north (Tipayamongkholgul et al., 2009). Using autoregressive Poisson models adjusted for seasonality and population density, a statistically significant association was identified between negative ENSO phases and dengue outbreaks, with time lags of up to 11 months (Tipayamongkholgul et al., 2009). The best correlations were observed when ENSO Indices were lagged by 4-6 months. Consistent with these findings, the best lag for SOI in our study was 5 months. On average, ENSO accounted for up to 22% of the monthly variability of cases in the northern provinces and 15% in the southern provinces (Tipayamongkholgul et al., 2009). In our study, the models incorporating SOI and climate parameters accounted for 53.7% of the variance in the data.

A longitudinal study integrating data from Puerto Rico, Mexico, and Thailand evaluated the association between ENSO, local climatic variables, and dengue incidence through time-frequency domain coherence analysis (wavelet analysis) (Johansson et al., 2009). In Puerto Rico, a transient relationship was identified among ENSO, temperature, and dengue incidence; however, on multi-year scales, only rainfall was significantly correlated with reported cases. However, the El Niño phenomenon, which generates droughts and higher temperatures in this country, is an important predictor of arbovirus disease outbreaks up to a year in advance (Barrera et al., 2023). In Thailand, both temperature and precipitation were associated with ENSO; however, the link between precipitation and dengue cases was non-stationary (Johansson et al., 2009). In contrast, in Mexico, no significant associations were found between ENSO, climate, and dengue on a multi-year scale. A study in Brazil, using a hierarchical spatio-temporal model grounded on a Bayesian framework, explored the relationship between ENSO, seasonal weather fluctuations, and the larval index, while adjusting for population density and wealth inequalities (Pirani, M. 2024). El Niño events, higher seasonal rainfall, and temperatures above 23.3 °C were all strongly associated with increases in the Aedes aegypti larval index. Mosquito proliferation was especially pronounced in socially vulnerable municipalities, highlighting the role of environmental and social factors in vector abundance. A similar study was conducted in Peru, adopting a Bayesian hierarchical modeling approach to address spatial correlation and B-splines with four knots per year to address temporal correlation, to analyze ENSO and climate parameters (Dostal, T., 2022). Temperature and ENSO significantly increased the risk of dengue outbreaks, especially in summer and in some regions. Rainfall showed no strong independent effect. These findings suggest that local climatic conditions, urbanization patterns, and other contextual factors may modulate ENSO’s influence on dengue epidemiology. A study mapped the spatial association between ENSO activity (SOI) and dengue fever incidence across the Americas from 1995–2004, including the major 1997–1998 El Niño event (Ferreira, M. C., 2014). The authors analyzed standardized SOI and annual dengue incidence using regression and spatial interpolation to identify geographic patterns. Results show stronger ENSO–dengue coupling in Mexico, Central America, the northern Caribbean, and northern South America, with epidemic centers shifting southward and several countries exhibiting higher dengue incidence during El Niño phases. Our results extend these findings to Argentina. Recent findings indicate that the intensity of the El Niño phenomenon is likely to increase as climate change progresses (Cole, J. E., 2026), underscoring the need to better characterize its impact on dengue and other mosquito-borne infectious diseases.

Higher temperatures, associated with El Niño events, have been shown to increase the incidence of dengue, possibly by favoring viral replication and vector behavior (Ferreira D. H. S., 2022). Similar results have been observed in a study in 14 island nations in the South Pacific (Hales et al, 1999). In line with these findings, we observed a higher dengue risk at temperatures between approximately 20°C and 30°C during the El Niño and neutral phases, with less consistent effects during the La Niña phase (Fig. 2A). Above 30°C, dengue risk dropped significantly, which may be related to lower mosquito survival in high-temperature regions (Couper, L. I. 2025). Studies in Paraguay have also observed a similar U-shaped relationship (Gómez Gómez et al, 2022). Humidity, which is also a significant factor in boosting mosquito survival (Couper, L. I. 2025), exhibited different patterns across ENSO phases. During the La Niña (dry, cold) phase, humidity was a strong conditioner of dengue risk. During neutral and El Niño phases, the effect was less marked. Rainfall was not associated with dengue risk, in concordance with previous observations (Dostal, T. 2022).

Recent evidence suggests that non-climatic mechanisms may also contribute to dengue outbreaks. While our results do not permit mechanistic inferences, it is worth reviewing some of these non-climatic mechanisms, as they might help explain some findings. Prolonged ENSO events can affect drinking water availability and the deterioration of sanitation infrastructure, which can force vulnerable communities to store water under precarious conditions, thereby contributing to the proliferation of Aedes aegypti breeding sites (Stewart Ibarra et al., 2013). This might account for the SOI effect observed in this study after controlling for covariates.

Population displacement might also be a relevant factor. A study of the South Pacific islands detected synchrony in dengue outbreaks between neighboring countries, even in the absence of direct local climate correlations (Stewart Ibarra et al., 2013). The authors hypothesized that the virus could spread between islands via flows of infected people, facilitated by regional maritime and air transport networks. In urban areas with high territorial connectivity, such as many cities in Argentina, movement between provinces and/or borders could accelerate the introduction of the virus and/or vector into new areas, even without substantial changes in the local climate.

Transfrontier population movements may also help explain the results of our spatial analysis, which revealed a residual higher incidence of dengue in the northern states that border Paraguay and Brazil (Gómez Gómez et al., 2022; Lenharo, M. 2024). Brazil, facing the worst dengue fever situation globally, reported over 3 million cases in 2023 and over 1 million cases in the first part of 2024 (Lenharo, M. 2024). Dengue has been endemic in Paraguay since 2009, with all four dengue virus serotypes circulating since 2011 (Gómez Gómez et al., 2022). Conversely, the high residual risk in Argentina’s southern provinces is harder to explain. These are cold, dry regions where the common Aedes aegypti mosquito is not expected to survive. However, acclimated A. aegypti (Kramer, I. M., 2021) and cold-resistant strains of A. albopictus (Tippelt, L., 2020) may be responsible for dengue outbreaks in these regions. It must be noted that these considerations have not been directly addressed in our models. Therefore, they are purely speculative. More research is needed to test these hypotheses.

The findings presented herein must be interpreted within the framework of the limitations inherent in the methodological design used. This study adopts an ecological observational approach, in which the unit of analysis is the department, district, or commune of Argentina. Although this approach is adequate for identifying spatio-temporal patterns and generating hypotheses at the population level, it presents limitations when it comes to establishing causal relationships or extrapolating the results at the individual level, which is known as the ecological fallacy, a frequent limitation in this type of analysis (Villeneuve, P. J.,2022).

Likewise, although official sources and high-resolution georeferenced databases were utilized, some indicators may exhibit measurement errors, temporal inconsistencies, or underreporting, especially in regions with limited epidemiological surveillance coverage or reduced availability of up-to-date sociodemographic data. Additionally, it is worth noting that Argentina exhibits high structural heterogeneity, characterized by the coexistence of highly urbanized areas with extensive rural regions of low population density. This aspect poses challenges for homogenization and comparison across jurisdictions.

On the other hand, although a wide range of environmental, climatic, and structural variables has been incorporated, the presence of unobserved factors or complex interactions not captured by the models used cannot be ruled out. Variables such as prevention practices, vector control campaigns, the circulation of different serotypes of the virus, human mobility, and socioeconomic conditions significantly impact the local dynamics of dengue (Marti et al., 2020). Still, they may be difficult to incorporate into ecological-scale models. Extreme climatic events, such as periods of very intense rainfall or extreme heat, may also contribute to dengue fever outbreaks (Lopez et al., 2025) and should therefore be considered in future studies.

Another important limitation is the lack of standardized entomological surveillance data. Mosquito abundance is a key intermediate determinant linking climatic variability to dengue transmission. However, nationwide data on Aedes aegypti population indices with sufficient spatial and temporal coverage were unavailable for the entire 2018–2025 study period. Consequently, our analyses cannot evaluate whether the observed associations between SOI and dengue incidence are mediated by changes in mosquito abundance. Future studies integrating entomological surveillance with climatic and epidemiological data could help elucidate this causal pathway.

Finally, although modeling and validation strategies have been applied, the interpretation of the results must take into account that this is a descriptive and exploratory study, whose main objective is to generate knowledge about spatiotemporal patterns and possible population determinants of dengue to predict likely epidemiological outbreaks, rather than to identify definitive causal relationships.

## 5. CONCLUSION

In this ecological study extending from 2018 to 2024, we found an association between ENSO and dengue outbreaks in Argentina. Negative SOI scores, which are related to the El Niño phenomenon, were associated with dengue outbreaks after adjusting for covariates. Monthly temperatures between 20° and 30° Celsius during the El Niño and neutral phases, humidity > 50% during the La Niña phase, and population density were also significant predictors of higher dengue relative risk. Our results also suggest the presence of unobserved factors influencing dengue risk. Outbreak modeling strategies, which often drive health policy, may benefit from incorporating ENSO measures when they are relevant. Improved modeling may open new possibilities for developing early warnings to predict dengue outbreaks.

## Author’s roles

AE: Conceptualization, Formal analysis, Writing – review and editing

CEL: Methodology, Writing – original draft

MAM: Methodology, Writing – original draft

GL: Methodology, Writing – original draft

LMML: Methodology, Writing – original draft

CMP: Methodology, Writing – original draft

SDOL: Methodology, Writing – original draft

OAB: Methodology, Writing – original draft

MB: Conceptualization, Writing – review and editing

SPLL: Conceptualization, Funding acquisition, Supervision, Writing – review and editing

## Data Availability

All data produced in the present study are available upon reasonable request to the authors

